# Spatial Visualization of Cluster-Specific COVID-19 Transmission Network in South Korea During the Early Epidemic Phase

**DOI:** 10.1101/2020.03.18.20038638

**Authors:** James Yeongjun Park

## Abstract

**Background:** Coronavirus disease 2019 (COVID-19) has been rapidly spreading throughout China and other countries including South Korea. As of March 12, 2020, a total number of 7,869 cases and 66 deaths had been documented in South Korea. Although the first confirmed case in South Korea was identified on January 20, 2020, the number of confirmed cases showed a rapid growth on February 19, 2020 with a total number of 1,261 cases with 12 deaths based on the Korea Centers for Disease Control and Prevention (KCDC).

**Method:** Using the data of confirmed cases of COVID-19 in South Korea that are publicly available from the KCDC, this paper aims to create spatial visualizations of COVID-19 transmission between January 20, 2020 and February 19, 2020.

**Results:** Using spatial visualization, this paper identified two early transmission clusters in South Korea (Daegu cluster and capital area cluster). Using a degree-weighted centrality measure, this paper proposes potential super-spreaders of the virus in the visualized clusters.

**Conclusion:** Compared to various epidemiological measures such as the basic reproduction number, spatial visualizations of the cluster-specific transmission networks and the proposed centrality measure may be more useful to characterize super-spreaders and the spread of the virus especially in the early epidemic phase.

## Introduction

The first pneumonia cases of unknown origin were identified in Wuhan in early December 2019.^1^ Since then, coronavirus disease 2019 (COVID-19) has been rapidly spreading throughout China and other countries including South Korea. As of March 17, 2020, a total of 198,181 laboratory-confirmed cases had been documented globally with 7,965 deaths. The World Health Organization (WHO) has declared COVID-19 an international public health concern.^2^ The confirmed patients in South Korea had either visited or came from China. Secondary and tertiary transmissions have occurred since then, which have led to an accelerating rate of transmission in South Korea. As of March 17, 2020, a total number of 8,320 cases and 81 deaths had been documented in South Korea.

## Method

With the launch of COVID-19 data hub, officials from the White House and other national organizations issued a call to action for researchers in a multitude of disciplines such as computer science, epidemiology, economics, and statistics. Open access data such as epidemiological data, interactive web-based dashboards, and descriptive statistics have informed many about the current state of the pandemic.^3,4^ With a concomitant effort to combat the virus and to better understand virus etiologies, Korea Centers for Disease Control and Prevention (KCDC), an organization under the South Korean Ministry of Welfare and Health, has made many datasets available online that are unique to COVID-19 confirmed South Korea cases.^5^ The datasets only include confirmed COVID-19 patients with unique numeric patient identifiers, geographical data, and infection information if available. In an epidemiological dataset, they released the region of the affected patient, the identifier of the person who infected the patient, and the number of contacts with other people. The aim of this report is to create spatial visualizations of early COVID-19 transmission networks in South Korea using these data, which may indicate transmission patterns for each network.

The time series data of COVID-19 status in South Korea is analyzed to provide updated statistics. Using a spatial visualization of confirmed patients during an early epidemic phase, two major clusters are identified. As of March 12, 7,869 positive cases had been documented in South Korea, and 70 positive cases have information of the identifiers of who infected them. Although the first confirmed case in South Korea was identified on January 20, 2020, the number of confirmed cases showed a rapid growth on February 19, 2020 with a total number of 1,261 cases with 12 deaths based on the KCDC.^6^

As of March, newly reported cases in South Korea show that the numbers of positive cases and deaths seem to be declining and new cases remain within known clusters. Therefore, identifying early clusters and examining the confirmed cases in these early clusters, from January 20, 2020 to February 19, 2020 are crucial because these clusters remain the longest lasting sources of transmission. Out of 70 patients, only a subset of patients infected from confirmed cases from an early epidemic phase (January 20, 2020 to February 19, 2020) is used to create the network from the epidemiological data to further visualize the transmission networks of these two clusters. All the analysis and visualizations are performed using the ggplot2 software in R as well as Cytoscape.^7,8^

## Results

The time series data contains both overall statistics such as the number of tests as well as geographical data within South Korea from January 20, 2020 to March 12, 2020. Figure 1 shows the time series data of the cumulative COVID-19 statistics from January 20, 2020 to March 12, 2020.

**Figure 1.**
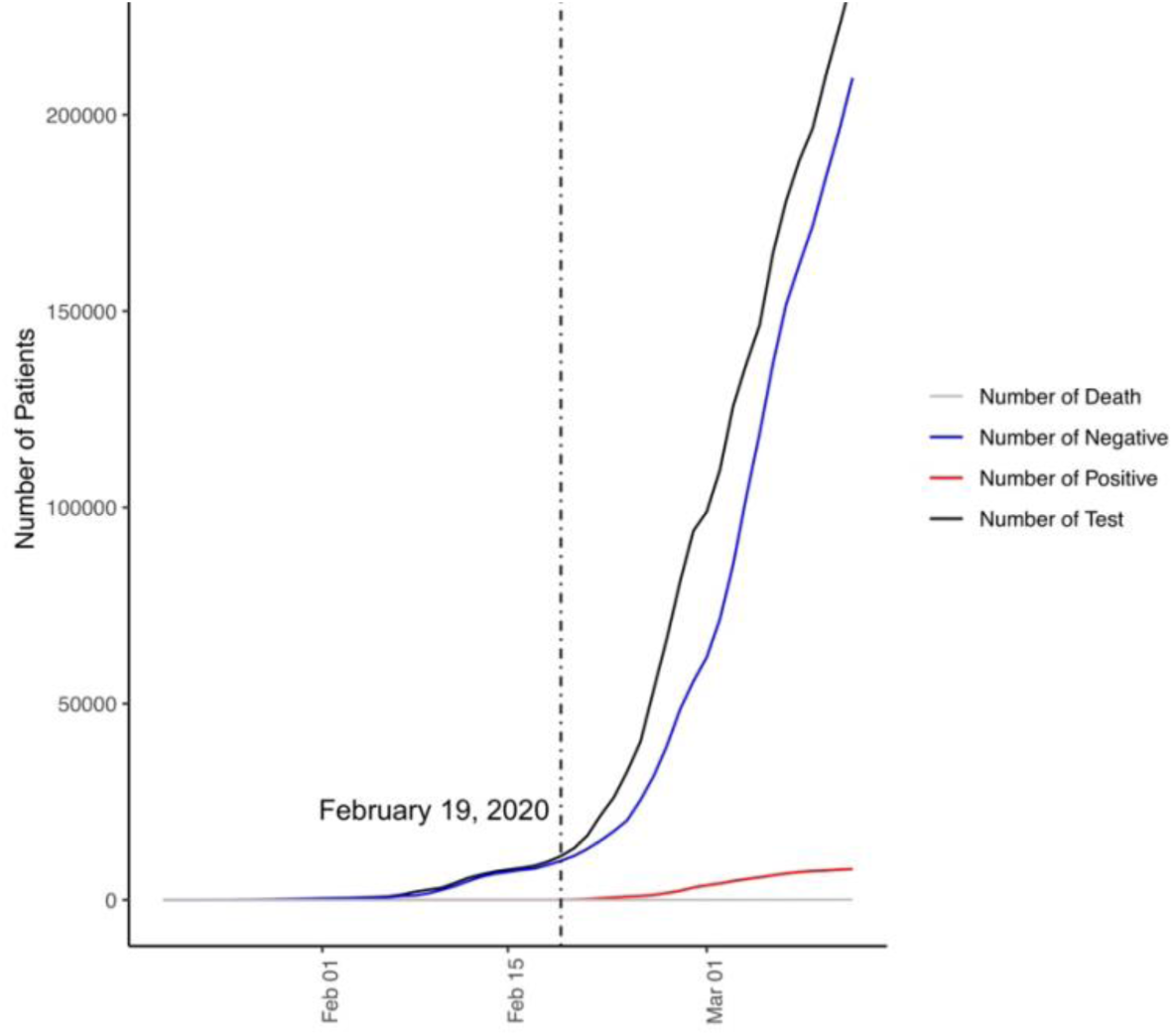
Time series data of the cumulative statistics of COVID-19 in South Korea from January 20, 2020 to March 12, 2020.

On January 20, 2020, there was one confirmed case from one test. On March 12, there were a total number of 234,998 tests where 209,402 patients were tested negative. 7,869 patients were tested negative as of March 12, 2020 of which 66 patients died. Over 53 days, on average, 3867 patients were tested for the virus of which 3133 patients were tested negative and 138 patients were tested positive each day. Since early February, there has been an exponential increase in the number of tested cases where most of them were tested negative. Out of 67 patients that were confirmed positive in South Korea as of February 19, 2020, the geographical data of 56 them are available, which allows a visualization of their route patterns. Figure 2 depicts the spatial distribution of the two large clusters of COVID-19 as of February 19th, 2020. The top left cluster corresponds to the capital area, and the bottom right cluster corresponds to Daegu, a city of 2.5 million people, approximately 150 miles away from the capital area.

**Figure 2.**
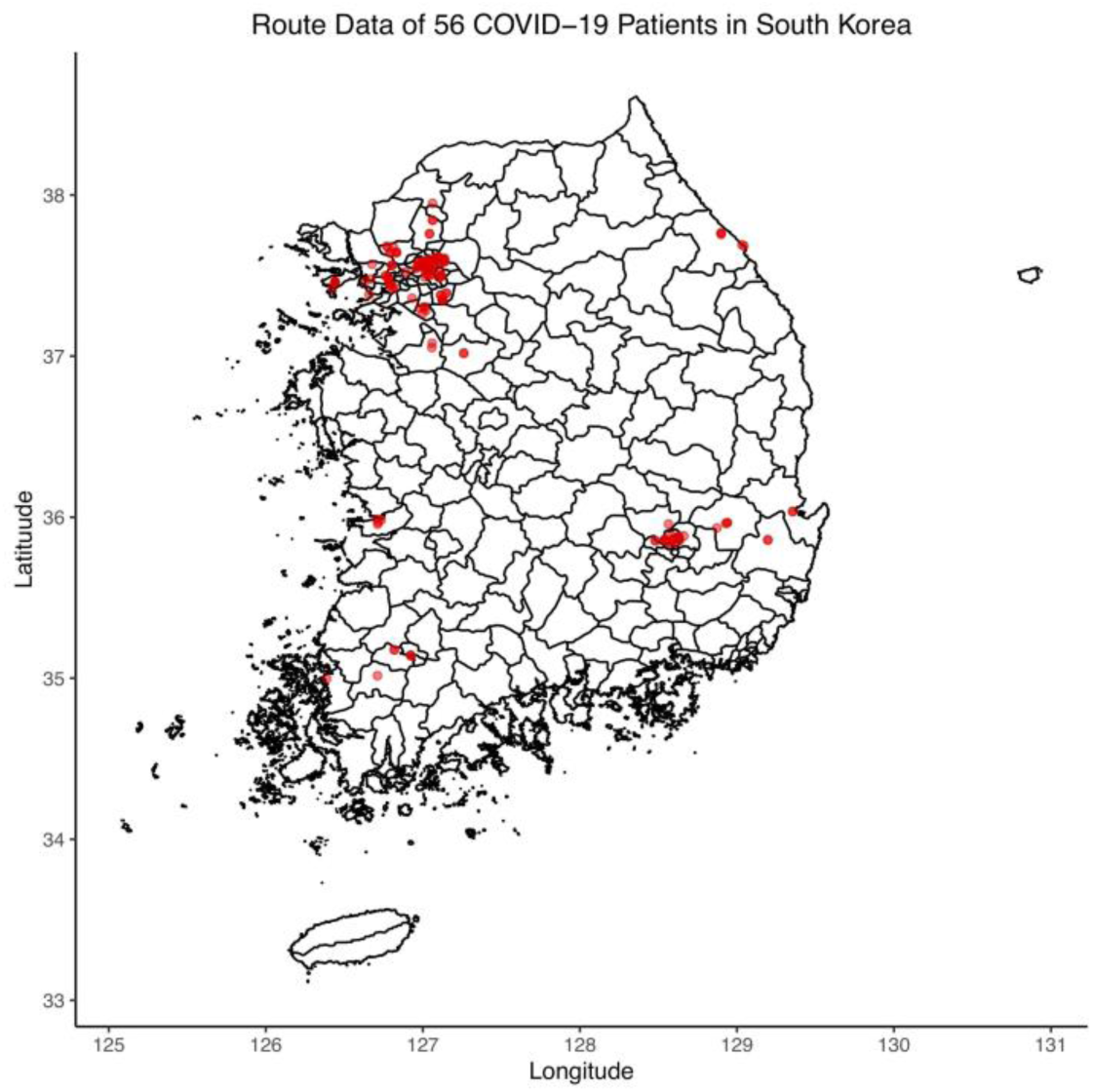
Route data of 56 COVID-19 confirmed cases in South Korea from January 20th, 2020 to February 19th, 2020

Figure 3 shows the transmission network of the capital area cluster. The numeric node numbers indicate unique patient identifiers and *n*^*th*^ COVID-19 confirmed cases in South Korea. The first case (the 3rd case in the nation) in the capital cluster was diagnosed positive on January 26, 2020. This patient had physical contact with 16 different individuals, travelled from Wuhan, China, and transmitted SARS-CoV-2 to the 6th and 28th confirmed cases in Korea. These two cases have further resulted in subsequential (i.e., secondary) positive cases in the capital area. In this graph, the 6^th^ case made physical contact with 17 different individuals and resulted in four new cases. The 6^th^ case is unlikely to be a super-spreader given a low number of physical contacts with other individuals before being treated. Although this cluster represents the largest connected component from the entire visualized infection network, it is reported that no further cases have been added in this cluster since February 21, 2020.^9^

**Figure 3.**
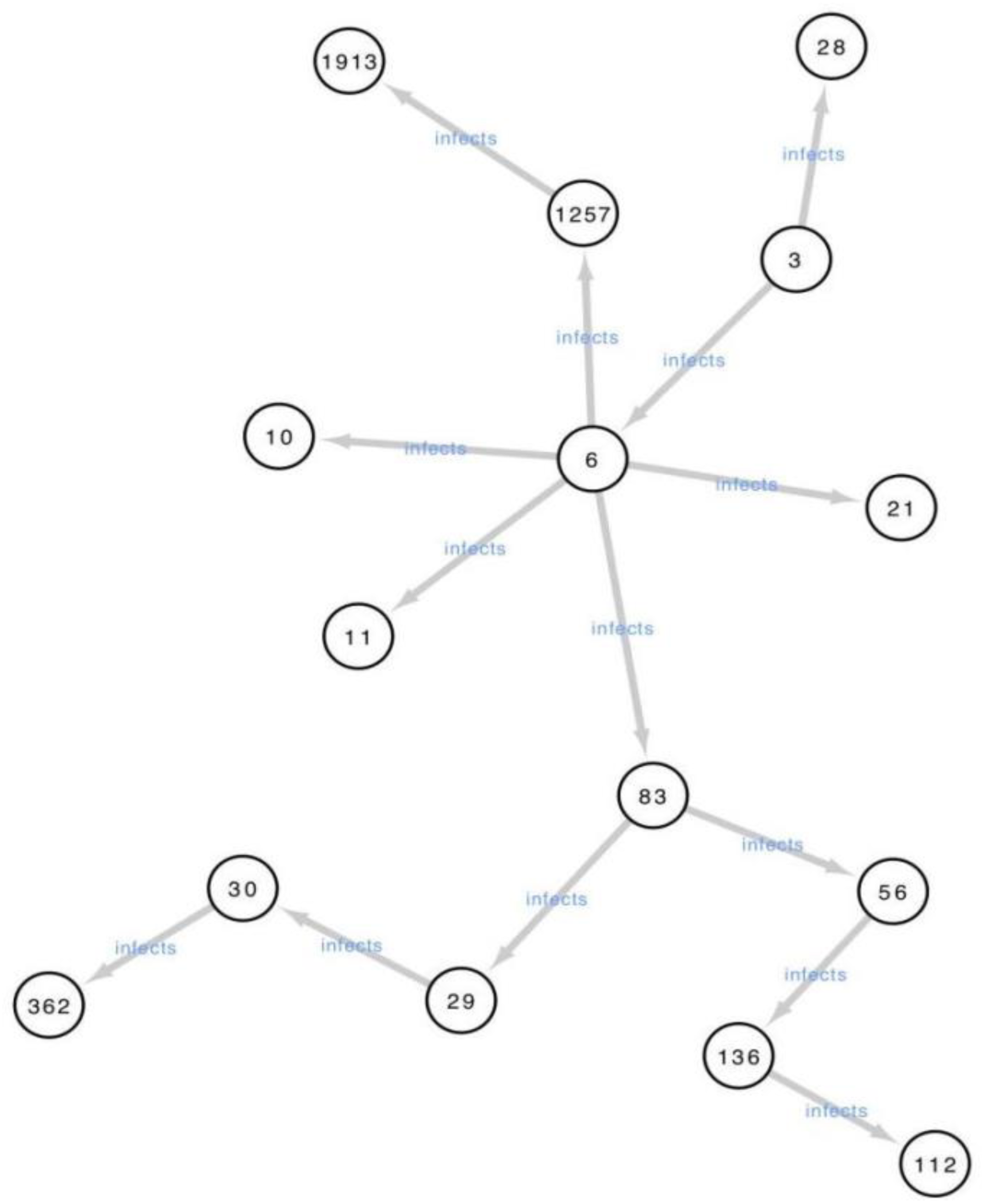
The transmission network of the capital area cluster composed of 15 different nodes

The epicenter of the South Korean COVID-19 outbreak has been identified in Daegu, associated with the Shin-Cheonji Church of Jesus. In this cluster, the first confirmed case—the 31^st^ patient in the cluster—was confirmed positive on February 18, 2020. This person had physical contact with an estimated number of 1,160 individuals in different places such as Shin-Cheonji Church of Jesus and the hospital in Cheong-do. Figure 4 clearly demonstrates how the first patient in this cluster infected eight individuals. This epicenter has been attributed as one of the major events that has led to at least 40 secondary cases in the city of Daegu and almost a half of the country’s confirmed cases were linked to this cluster in late February.^8^ However, secondary infection information was not available in the dataset, which does not allow visualization of a bigger network. Regardless, the Daegu network shows what is known as a super-spreader who contacted 1,160 different patients and started the transmission cluster in Daegu.

**Figure 4.**
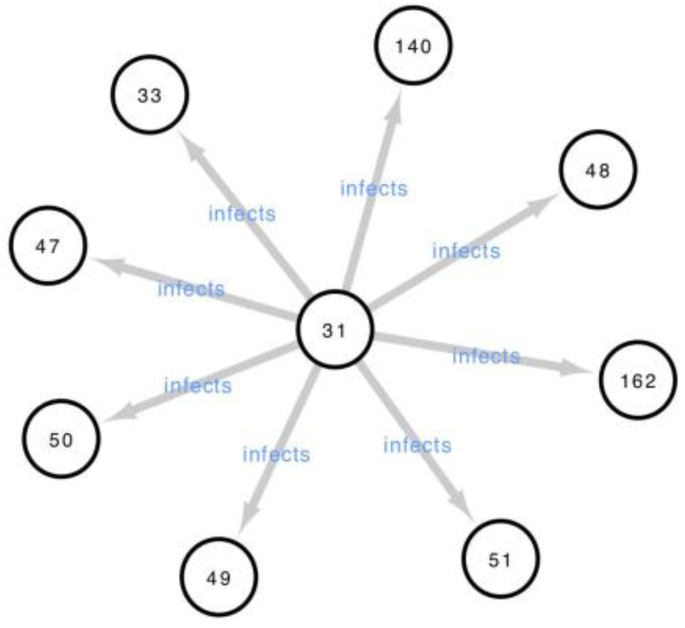
The transmission network of the Daegu cluster composed of 9 different nodes with one super node

These two clusters depict very different infection information through different topologies. Therefore, visualization of these clusters may be helpful to understand the transmission in addition to epidemiological measures like the reproduction number. A recent paper estimated the reproduction number and the outbreak size of the COVID-19 in South Korea using the deterministic mathematical model. The estimated *R*_0_ in the national level was 0.55 while the estimate in Daegu was between 3.47 - 3.54.^10^ Another estimate of *R*_0_ in South Korea was 1.5 (95% CI: 1.4 - 1.6), calculated using the generalized growth model.^9^ These estimates are dynamic, and global estimates of the reproduction numbers may not be easily interpretable for different region- and group-specific transmission clusters. There are also a number of different mathematical models that can be used to calculate the reproduction number under different probability distributions, which further complicates the interpretability. Instead, visualizing the transmission networks could be useful to understand the spread of the virus.

Although it may be clear that the 31^st^ case in the Daegu cluster is a super-spreader, it is uncertain who is the super-spreader in the capital area cluster. Out of 15 distinct cases in the network, nine cases have reported degrees in the dataset (number of contacts), which allows the use of centrality algorithms to understand the role of particle nodes in a graph and their impact on this transmission network. *d*_*i*_ denotes the degree of the *i*^*th*^ case. Since six nodes are missing degrees, the population average degree is used to impute missing degree information. We define a population degree *d*_*p*_, which is calculated after imputing missing degrees with an assumption that every node in the network is independent of each other. Table 1 shows the number of degrees for each case before and after imputation.

**Table 1.** Number of degrees in the capital cluster before after imputation

| case number | degree | degree imputed |
| --- | --- | --- |
| 3 | 16 | 16 |
| 6 | 17 | 17 |
| 10 | 43 | 43 |
| 11 | 0 | 0 |
| 21 | 6 | 6 |
| 28 | 1 | 1 |
| 29 | 117 | 117 |
| 30 | 27 | 27 |
| 56 |  | 32 |
| 83 |  | 32 |
| 112 |  | 32 |
| 136 |  | 32 |
| 362 |  | 32 |
| 1257 |  | 32 |
| 1913 | 61 | 61 |

Betweenness centrality is another graph centrality measure that captures the influence of a node over the flow of information between every pair of nodes in the network with the assumption that information flows over the shortest paths between them. Between centrality *C*_*B*_(*n*) for a node *n* is defined as

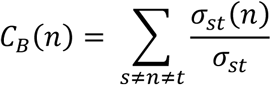

where *σ*_*st*_ is the number of shortest paths with edges *s* and *t* as their end edges while *σ*_*st*_(*n*) is the number of those shortest paths that include node *n*.^11^ We propose a degree-weighted betweenness centrality *C*_*BD*_(*n*), which prioritizes nodes with high degrees while penalizing them by *d*_*p*_

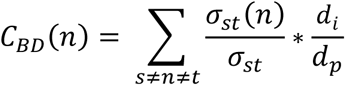

to capture the super-spreader in the capital area network. Table 2 shows the calculated metrics from the capital area network.

**Table 2.** Proposed centrality measures from the capital area network

| case number | $d_i$ | $C_B(n)$ | $C_{BD}(n)$ |
| --- | --- | --- | --- |
| 29 | 117 | 0.264 | 0.064 |
| 83 | 32 | 0.626 | 0.042 |
| 6 | 17 | 0.747 | 0.026 |
| 56 | 32 | 0.264 | 0.018 |
| 1257 | 32 | 0.143 | 0.01 |
| 136 | 32 | 0.143 | 0.01 |
| 30 | 27 | 0.143 | 0.008 |
| 3 | 16 | 0.143 | 0.005 |
| 28 | 1 | 0 | 0 |
| 21 | 6 | 0 | 0 |
| 11 | 0 | 0 | 0 |
| 10 | 43 | 0 | 0 |
| 1913 | 61 | 0 | 0 |
| 112 | 32 | 0 | 0 |
| 362 | 32 | 0 | 0 |

By looking at betweenness centrality only, the 6^th^ case who transmitted the virus to five distinct cases has the highest betweenness centrality. However, a degree-weighted measure indicates that the 29^th^ case with a much larger degree is the most central node in the network. This metric may be useful for small networks with limited information to identify super-spreaders in the early transmission networks.

## Discussion

What happened in China shows that quarantine, social distancing, and isolation of infected populations may be able to contain the epidemic.^12^ This is encouraging for the many countries where COVID-19 is beginning to spread. South Korea once had the fastest growing rate of infection outside of China. Korea’s confirmed cases have risen rapidly since the identification of the super node in the Daegu cluster since late February. Since then, the country has shown success in its mitigation efforts in both the number of newly confirmed cases and deaths. The majority of new cases originate from those original clusters, one of which is likely a super-spreader, which is suggested by the spatial network generated.

Similar observations were seen during the Middle East respiratory syndrome (MERS) in South Korea where the syndrome was spread rapidly by super-spreaders.^13^ Therefore, it is important to have a better understanding of these clusters during the early epidemic phase, and visualizing them may help us understand how the virus is being spread. Spatial networks can visualize early transmission clusters, and the proposed degree-weighted betweenness centrality measure can further help identify super-spreaders in the identified clusters, which may not only reduce the spread of the virus but may also help with policymaking such as enforced social distancing or quarantining.

## Data Availability

The datasets used in the manuscript are available publicly from the Korea Centers for Disease Control and Prevention.

